# The GA4GH Phenopacket schema: A computable representation of clinical data for precision medicine

**DOI:** 10.1101/2021.11.27.21266944

**Authors:** Julius O. B. Jacobsen, Michael Baudis, Gareth S. Baynam, Jacques S. Beckmann, Sergi Beltran, Tiffany J. Callahan, Christopher G. Chute, Mélanie Courtot, Daniel Danis, Olivier Elemento, Robert R. Freimuth, Michael A. Gargano, Tudor Groza, Ada Hamosh, Nomi L. Harris, Rajaram Kaliyaperumal, Aly Khalifa, Peter M. Krawitz, Sebastian Köhler, Brian J. Laraway, Heikki Lehväslaiho, Kent C. Lloyd, Leslie Matalonga, Julie A. McMurry, Alejandro Metke-Jimenez, Christopher J. Mungall, Monica C. Munoz-Torres, Soichi Ogishima, Anastasios Papakonstantinou, Davide Piscia, Nikolas Pontikos, Núria Queralt-Rosinach, Marco Roos, Paul N. Schofield, Anastasios Siapos, Damian Smedley, Lindsay D. Smith, Robin Steinhaus, Jagadish Chandrabose Sundaramurthi, Emilia M. Swietlik, Sylvia Thun, Nicole A. Vasilevsky, Alex H. Wagner, Jeremy L. Warner, Claus Weiland, Melissa A. Haendel, Peter N. Robinson

## Abstract

Despite great strides in the development and wide acceptance of standards for exchanging structured information about genomic variants, there is no corresponding standard for exchanging phenotypic data, and this has impeded the sharing of phenotypic information for computational analysis. Here, we introduce the Global Alliance for Genomics and Health (GA4GH) Phenopacket schema, which supports exchange of computable longitudinal case-level phenotypic information for diagnosis and research of all types of disease including Mendelian and complex genetic diseases, cancer, and infectious diseases. To support translational research, diagnostics, and personalized healthcare, phenopackets are designed to be used across a comprehensive landscape of applications including biobanks, databases and registries, clinical information systems such as Electronic Health Records, genomic matchmaking, diagnostic laboratories, and computational tools. The Phenopacket schema is a freely available, community-driven standard that streamlines exchange and systematic use of phenotypic data and will facilitate sophisticated computational analysis of both clinical and genomic information to help improve our understanding of diseases and our ability to manage them.

## Introduction

Despite the clinical importance of phenotypic features (signs, symptoms, laboratory and imaging findings, results of physiological tests, etc.), exchanging them in conjunction with genomic variation is often overlooked or even neglected. Although great progress has been made in exchange formats for sequence and variation data such as the Variant Call Format (VCF),^1^ complementary standards for phenotypic and other clinical data have lagged far behind. In the clinical domain, a significant amount of work has been dedicated to the development of computational phenotypes.^2^ Traditionally, these approaches have largely relied on rule-based methods and large sources of clinical data to identify cohorts of patients with or without a specific disease.^3–6^ Unlike the aforementioned genomic standards, these approaches were not developed to model patient-level phenomena nor facilitate complex within and between patient comparisons. For example, one cannot readily compare an observed phenotypic profile against clinical databases as one can with genomic sequences using tools such as BLAST,^2^ partially because the lack of standards has impeded the development of algorithms and software for computational phenotype analysis. Another limitation is that phenotypic abnormalities of individuals are currently described in diverse places in multiple formats:^7^ scientific publications, biomedical databases, health records, patient health forums,^8^ and social media.^9^ The development of a reliable and practical standard for the documentation and exchange of clinical phenotypes is then critically necessary. A phenotype exchange standard is also *timely* for two reasons: firstly, there is increasing consensus about best practices for secure phenotypic data sharing and secondly, there have been significant advances in analytical methods to make effective use of computationally encoded phenotypic data.^10^

Deep phenotyping is a process whereby a multitude of morphological, biochemical, physiological, or behavioral attributes of each patient are collected by traditional clinical examinations and increasingly through advanced technologies such as imaging.^11,12^ Many diagnostic tools now leverage phenotype-driven differential diagnostic support on the basis of ontologies such as the Human Phenotype Ontology (HPO).^13–30^ Ontologies are systematic formal representations of knowledge that can be used to integrate and analyze large amounts of heterogeneous data by defining entities and concepts such as genetic variation, clinical findings, and diseases as well as the relationships between these concepts in a way that allows computational logical reasoning.^10^ Ontology-driven algorithms have been transformative for rare disease diagnosis.^30,31^ While the HPO^32^ offers over 16,000 terms to describe phenotypic abnormalities (symptoms, signs, laboratory abnormalities, behavioral manifestations, imaging findings, etc.), it does not itself provide a framework for exchanging information about the phenotypic abnormalities comparable to the way VCF provides a framework for exchanging information about all of the variants found in an exome or genome sequence. Such a framework would greatly facilitate precision medicine and precision public health.^10^

One challenge is that computational phenotype analysis is still poorly connected with the Electronic Health Record (EHR) and also that EHRs are not standardized across institutions or countries^33^. To enable precision medicine, standards and tools are needed to improve machine readable phenotypic characterization of patients beyond current standard EHR billing and clinical encounter data capture; EHR problem lists can only partly address this problem. To study disease trajectories and to optimize care plans for patients there is a need to identify correlations and changes in phenotypic characteristics with other data modalities such as imaging, patient-reported data including mobile health, ‘omics data, etc. Standardization of phenotype data is necessary to gather together the sample sizes of hundreds of thousands of patients from widely distributed sources, as will be required to assess the clinical relevance of (rare) genetic variants in Mendelian and common diseases with sufficient statistical support.^34–36^ Many complex and infectious diseases would also benefit from a robust, computational representation of phenotypic characteristics and their temporal progression in association with genomic, other ‘omics, environmental, mobile health data, etc.^10^ Further, increasingly detailed phenotypes are required to improve our understanding of how specific variants can predict patient phenotypic sub-groups, complications, progression rates, and/or response to therapy. Scalable deep phenotyping will be important for both rare and complex diseases within learning health systems.^37,38^

The Global Alliance for Genomics and Health (GA4GH) is developing a suite of coordinated standards for genomics for healthcare.^39^ The Phenopacket is a new GA4GH standard for sharing disease and phenotype information. To build the standard, requirements and specifications were established through a community effort. The standard underwent a rigorous peer review and product approval process and was consequently promoted as a GA4GH standard. Version 1 of the GA4GH standard was released in 2019 to elicit feedback from the community. Version 2 was developed on the basis of this feedback and is described here. A Phenopacket characterizes an individual person or biosample, linking that individual to detailed phenotypic descriptions, genetic information, diagnoses, and treatments (Figure 1). The Phenopacket schema enables comparison of sets of phenotypic attributes from individual patients. Such comparisons can aid in diagnosis and facilitate patient classification and stratification for identifying new diseases and treatments.^10^ The Phenopacket schema is designed to support interoperability between the people, organizations, and systems that comprise the worldwide effort to address human disease and biological understanding. These partners include clinical laboratories, authors, journals, clinicians, data repositories, patient registries, EHRs, and knowledge bases; the structure of the information in a phenopacket was designed for integration within these distributed contexts. Increasing the volume of computable data across a diversity of systems will support global computational disease analysis by integrating genotype, phenotype, and other multi-modal data for precision health applications.

**Figure 1.**
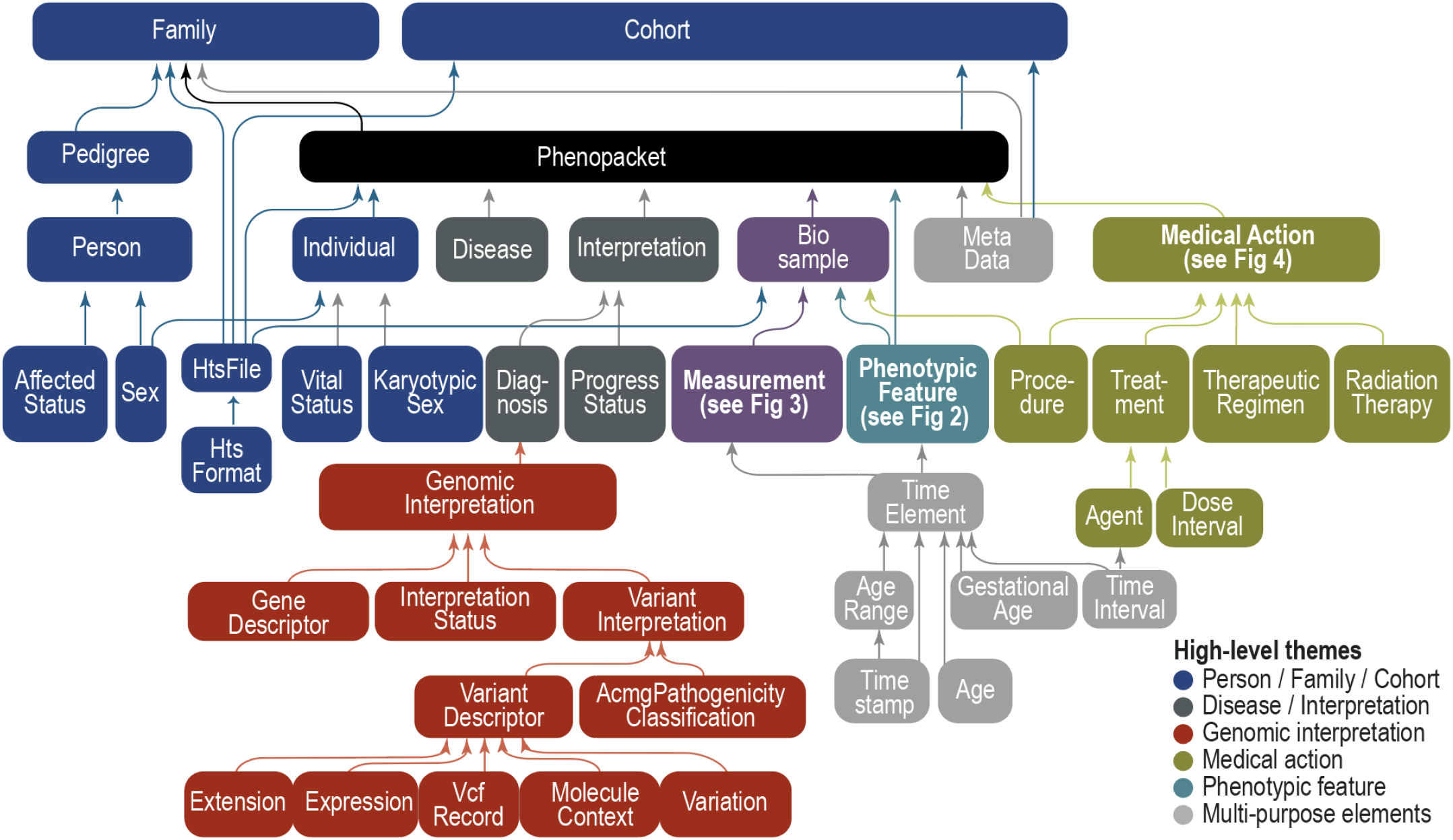
Phenopacket schema overview. The GA4GH Phenopacket schema consists of several optional elements, each of which contains information about a certain topic such as phenotype, variant, pedigree, etc. One element can contain other elements, which allows a hierarchical representation of data. For instance, the Phenopacket contains elements of type Individual, PhenotypicFeature, Biosample, and so on. Individual elements can therefore be regarded as **building blocks** that are combined to create larger structures. Colors represent the major themes of elements within the schema. See Figures 2, 3, and 4 for specifications of the PhenotypicFeature, Measurement, and Medical Action elements.

## Results

The GA4GH Phenopacket Schema is a set of rules that specify the organization of data as a blueprint for constructing a phenopacket, which is a structured representation of an individual’s medically relevant data, providing a computable case report of either a single medical encounter or a time course that can represent the entire medical history of an individual. It includes information such as age (which can be represented in multiple ways including ranges to protect privacy), sex, and gender as well as existing disease diagnoses. Almost all elements of a phenopacket are optional. A simple phenopacket contains only information about the individual and a list of phenotypic features and is sufficient for use cases of Mendelian disease genomic diagnostics. More comprehensive phenopackets contain data about biosamples and treatment associated with an individual patient along with phenotypic features and can be appropriate for use in clinical management beyond diagnosis, such as treatment selection.

### Schema overview

Currently, a very wide range of *ad hoc* database schemas are used to represent clinical data for specific research projects, and numerous different ontologies are used to represent clinical entities; for instance, diseases can be represented by terms from ontologies and terminologies including Mondo, OMIM, Orphanet, NCIT, ICD, and many others.^40^ Therefore, the Phenopacket schema is intentionally flexible to facilitate wide adoption and to increase the value of the network of systems sharing phenopackets for computational use. The major goals for the Phenopacket schema include composability, traceability (data provenance), the FAIR principles (Findable, Accessible, Interoperable, and Reusable), and computability.^41–44^ Specifically, Phenopackets are designed to be both human and machine-interpretable, enabling computing operations and validation on the basis of defined relationships between diagnoses, lab measurements, and genotypic information; they also enable seamless transfer of data from a data source (e.g., a document describing the phenotypic information) to a data receiver (e.g., an application that receives and uses it). The standard supports several computational document formats such as JSON and YAML. Phenopackets require common and well-established ontologies, that is, logically defined hierarchies of terms that allow sophisticated algorithmic analysis over medically relevant abnormalities.^10^ The Phenopacket schema does not directly model -omics data in detail but does enable users to link a Phenopacket to files representing data from high-throughput screening techniques or to denote individual variants in several formats (see section on VRS and VRSATILE, below).^45^ The Phenopacket schema integrates the GA4GH Variant Representation Specification and is designed to be interoperable with other GA4GH standards including those for pedigree data. **Figure 1** highlights the key building blocks of the Phenopacket schema, some of which are described in further detail below.

## Elements of a Phenopacket

In the sections that follow, we describe a selection of the most important Phenopacket elements (see Figure 1). For the complete schema and detailed documentation for each element, please refer to the online documentation (see Web Resources).

### PhenotypicFeature

The *PhenotypicFeature* (**Fig. 2**) is the central element of the Phenopacket schema. A *PhenotypicFeature* can be used to describe each phenotypic feature (often, but not necessarily, clinical abnormalities) including signs and symptoms, laboratory findings, histopathology findings, imaging, electrophysiological results, etc., along with modifier and qualifier concepts. Sources of phenotypes include physicians, laboratories, and patients; as wearables become more ubiquitous and more capable, these too will be important sources of phenotypes and context. Each phenotypic feature is described using an ontology term. While the Phenopacket schema does not mandate which ontology to use, there are recommendations, such as the HPO for rare diseases and the National Cancer Institute Thesaurus (NCIT) for transmission of information about a cancer specimen such as pathological staging or more detailed information about histology or tumor markers.^46^ One can indicate whether a certain abnormality was excluded during the diagnostic process (e.g., whether a morphological cardiac defect was excluded by echocardiography), or use other optional HPO terms to denote the severity of the *PhenotypicFeature* or other modifiers that describe the frequency (e.g., number of occurrences of seizures per week), laterality (e.g., unilateral) or other pattern of a certain phenotypic feature in the patient being described. Finally, the onset (and if applicable the resolution) of specific features can be indicated. Further information on this and other elements is available in the online documentation (See Web Resources).

**Figure 2.**
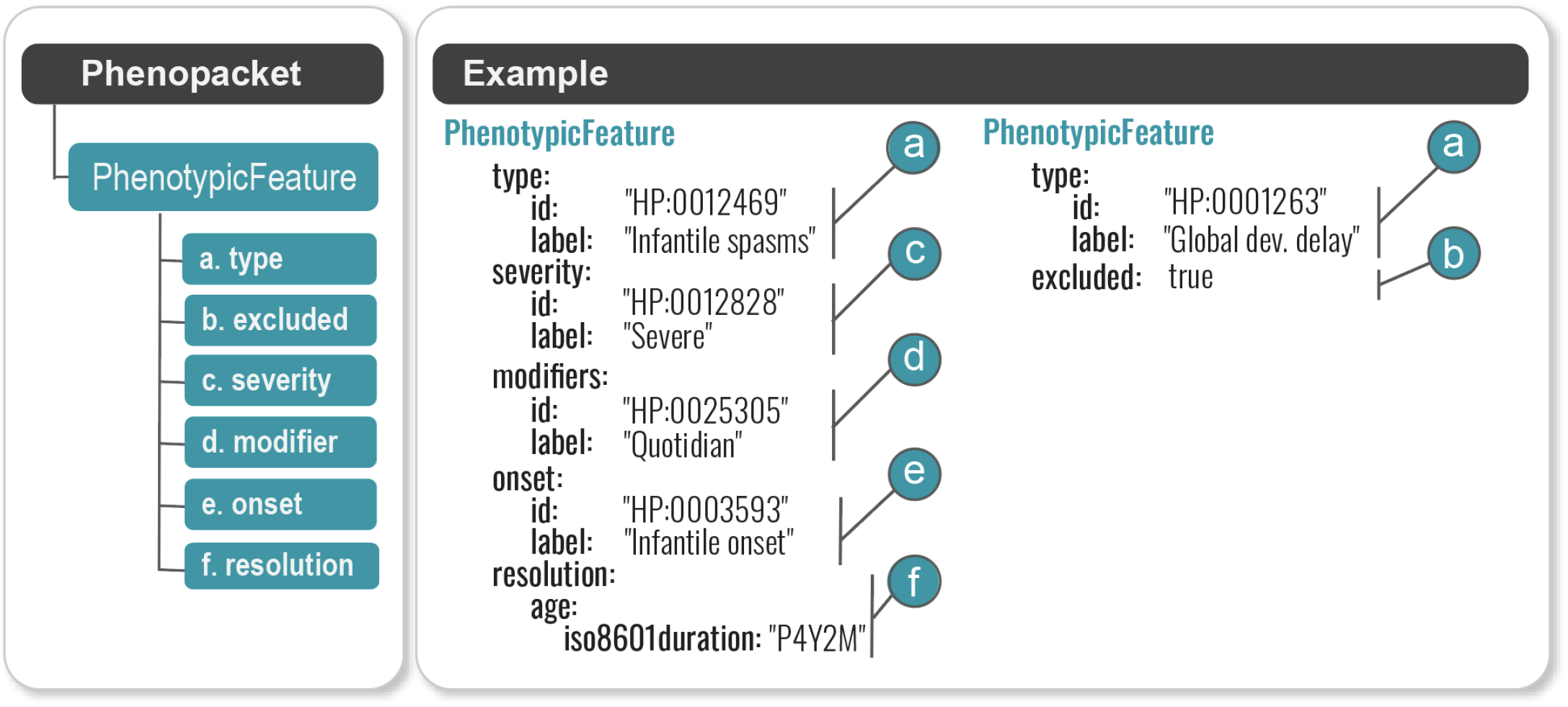
PhenotypicFeatures in a Phenopacket. A Phenopacket can contain information about an arbitrary number of phenotypic features observed in a single individual, each encoded using a PhenotypicFeature element. For medical use cases the subject will generally be a patient or a proband of a study, and the phenotypes will be abnormalities described by an ontology such as the HPO. Each phenotypic feature is defined by an HPO term (a), which is qualified as either present or absent (excluded) (b), with possible severity (c), modifiers (d), onset (e), and resolution (f). The example in the right panel shows a phenotypic feature, severe daily infantile spasms, which first occurred in infancy and resolved at age 4 years and 2 months, in a child without global developmental delay.

**Figure 3.**
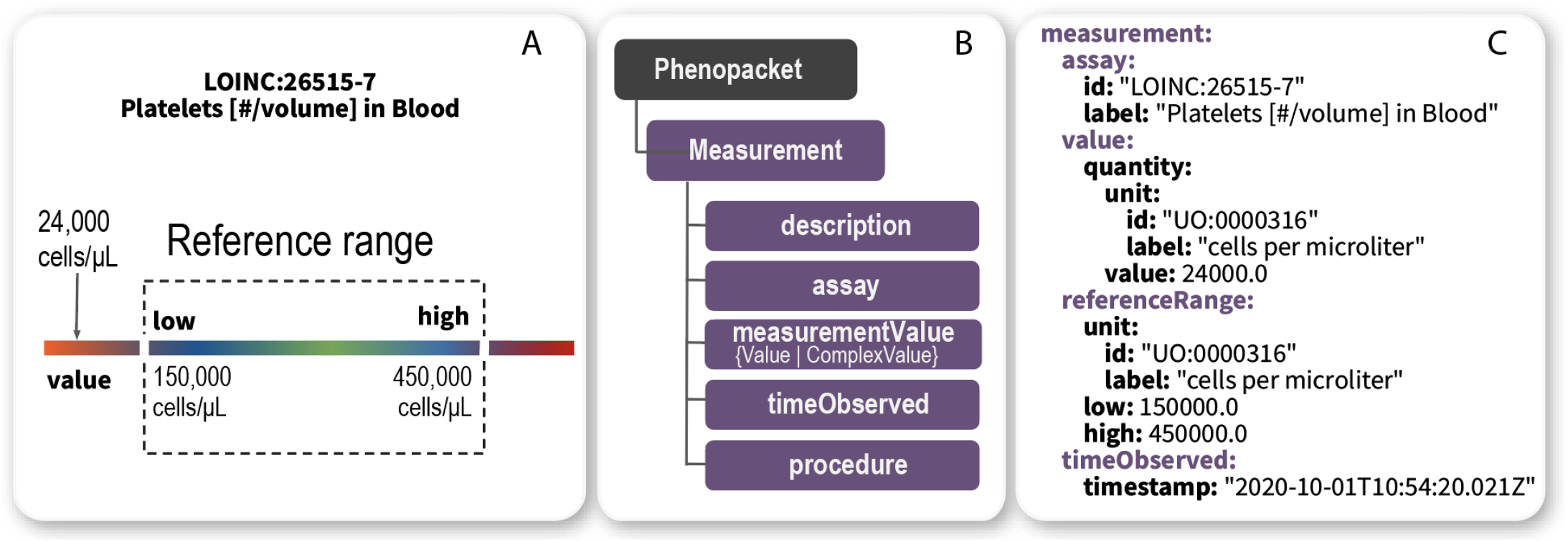
Example measurement of platelet count. Panel A: graphical representation of abnormal laboratory value outside of reference range. Panel B: relevant Phenopacket hierarchy. Panel C: part of a Measurement representing an abnormally low value for thrombocyte count. The reference range represents the range that was applied in the specific investigation and thus may reflect age or sex-specific values for some analytes.

**Figure 4.**
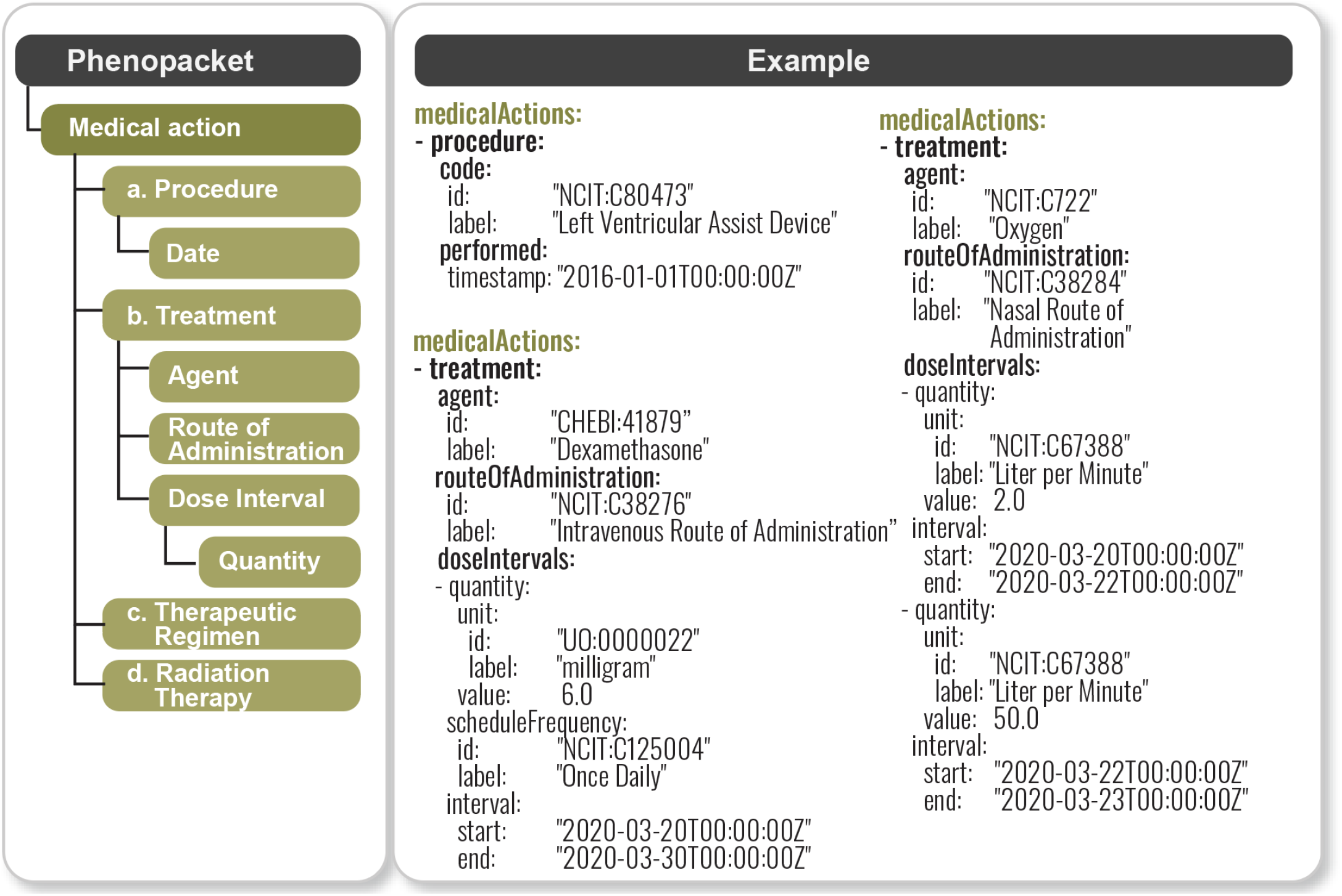
Phenopacket MedicalAction representation. Left panel: components of a *MedicalActio*n in a Phenopacket. Right panel: example using COVID medical actions: implantation of a left ventricular assist device in 2016 (this information from the past medical history is important as it represents a risk factor for severe COVID-19 infection), intravenous administration of dexamethasone, and provision of oxygen by nasal cannula at an initial dose of 2 liters per minute that was later increased to 50 liters a minute. In general, a *MedicalAction* consists of one of the four options (*Procedure, Treatment, TherapeuticRegimen*, or *RadiationTherapy*). A Phenopacket can include an arbitrary number of medical actions.

**Figure 5.**
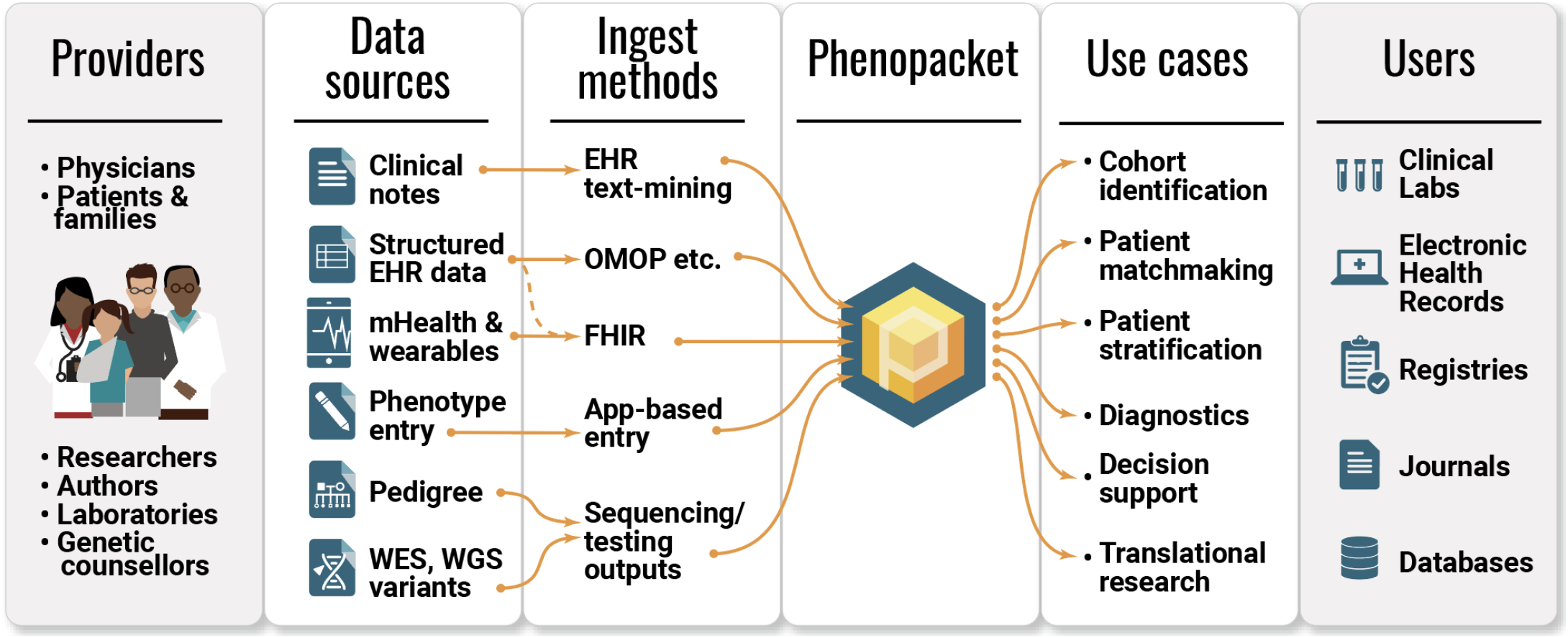
Phenotype data exchange in the biomedical ecosystem. Multiple providers of phenotypic data include patients and clinicians, via a variety of mechanisms including mHealth and the EHR. The Phenopacket schema acts as a common model that can capture data from many sources with a unified software representation and in turn can be used by multiple receivers of the phenotypic information, including journals, databases, registries, clinical laboratories. Phenopackets can support diverse users and use cases, including patient matchmaking services, diagnostics, and cohort identification.

### Measurement

The *Measurement* element is used to capture quantitative, ordinal (e.g., absent/present), or categorical measurements. For applications such as phenotype-driven genomic diagnostics of rare disease, qualitative representations of phenotypic abnormalities are appropriate, e.g., ‘Thrombocytopenia’ (HP:0001873). For other use cases, such as following the development of some parameter over time, the original quantitative values may be preferable, and can be represented using the *Measurement* element; for instance, 17,000 platelets per microliter. *Measurement* objects can be used to represent normal or abnormal measurements or to represent complex measurements with multiple components such as blood pressure.

### Biosample

A *Biosample* is a description of biological material obtained from the individual represented in the Phenopacket and used for phenotypic, genotypic, or other -omics analysis. For instance, a *Biosample* can represent a biopsy of an ‘infiltrating urothelial carcinoma’ (NCIT:C39853) taken from the urinary bladder wall (UBERON:0001256) by ‘radical cystoprostatectomy’ (NCIT:C51899) and found to be ‘stage II’ (NCIT:C28054) for which genome sequencing was performed, with a link to the VCF sequence variant file. If desired, *Biosample* can represent samples derived from other *Biosamples*, e.g., an RNA sample derived from a tumor. Interpretations of the genomic findings in a *Biosample*, if available, are represented in Interpretation elements (see below).

### *MedicalAction* including *Treatment*

For cancer,^47^ infectious disease^48^, rare disease,^49^ and many categories of common disease,^50^ precision-medicine approaches to treatment based on classification by genetic variants are rapidly gaining in importance. Consequently, clinical decision-making needs to integrate genomic research findings with structured information about treatments. The GA4GH Phenopacket schema includes a hierarchical representation of medical actions including medications, procedures, and other actions taken for clinical management. The *Treatment* element represents administration of a pharmaceutical agent, broadly defined as prescription and over-the-counter medicines, vaccines, and other therapeutic agents such as monoclonal antibodies or CAR T-cell-therapy.

### Interpretation

A Phenopacket can contain one or more *Interpretation* elements that specify interpretations of genomic findings. For instance, a report from a diagnostic laboratory about a variant interpreted to be causal for a certain Mendelian disease may be represented as an *Interpretation* element, as can a report of a potentially actionable somatic variant for which a targeted cancer therapy is available.

As a GA4GH standard, the Phenopacket schema integrates with and leverages other GA4GH standards when applicable. Subsequent to the release of the Phenopacket schema v1, the GA4GH Genomic Knowledge Standards Work Stream developed a precise and extensible standard for the representation of genomic variants: the Variation Representation Specification (VRS, pronounced “verse”). The Phenopacket schema v2 leverages the computational precision of VRS while maintaining the flexibility of describing variation using human-readable variant description formats (such as SPDI^51^ or HGVS^45^) through collaborative development and adoption of the VRS Added Tools for Interoperable Loquacious Exchange (VRSATILE, pronounced “versatile”; see Web Resources).

VRSATILE provides two primary object classes that are used in the Phenopacket schema: the *VariationDescriptor* and the *GeneDescriptor*. These descriptor classes allow for the extension of computationally precise concepts (e.g., VRS alleles, HGNC gene identifiers^52^) with common additional attributes for systems to describe these concepts (e.g., identifier cross-references, HGVS descriptions, gene symbols, and informative contexts such as variant zygosity). This collaborative framework bridges existing variant representation formats and implementations to the more computationally precise concepts enabled by VRS.

## Discussion

The VCF standard for storing genotyping data allowed a wide range of research groups to write software for analyzing such data.^1^ The GA4GH Phenopacket schema aspires to be similarly transformative in the landscape of genome analysis using phenotype data.

### Interoperability and integration of Phenopackets schema

#### Pedigree standards

Unambiguous and computable pedigree information is critical for performing family-based genomic analysis. Pedigree data is currently represented in heterogeneous formats, such as custom JSON formats that can challenge interoperability, and lowest-common-denominator formats like PED (pedigree format), which are often limited in the representation of more complex families. The Phenopacket schema currently represents pedigree information using a simple transformation of the common PED format, which allows for describing the basic parent-child relationships necessary for family-based genetic analysis. Use cases that require richer information, such as clinical pedigrees, patient-reported family history data, and risk assessment for hereditary cancer, will be supported by integrating the GA4GH Pedigree Standard currently in development.

#### GA4GH Beacon

While the initial GA4GH Beacon standard empowered federated search for specific genomic variants,^53^ recent and upcoming versions of Beacon extend its scope through the use of standardized phenotypic data in both queries and data transfer. Beacon v2 has implemented a flexible filters extension for specifying additional query parameters that use ontology terms specified through CURIEs, in line with the Phenopacket schema. For data delivery, in contrast to the Boolean (true/false) responses provided through the original Beacon protocol, Beacon v2 allows for rich data delivery with response content described in default or external schemas. Although the v2 default schemas already track the Phenopacket schema to a large extent (e.g., *Biosample*), Beacon providers can decide to directly deliver Phenopacket responses to Beacon queries. Here, the greater sensitivity of richer data to be transferred through federated Beacon networks adds emphasis to the careful use of the tiered access control to increase security.

#### EHR standards

Medical coding systems and clinical exchange standards have not to date included rich phenotypic descriptions, as they are largely focused on supporting billing and clinical encounter documentation, rather than the documenting and sharing of the biologically relevant phenotypic information needed for computational use, mechanism discovery, and precision classification. From a clinical perspective, the integration of a standard for phenotypic description and exchange into and out of EHRs would improve disease diagnosis and management, especially for genomic health and precision medicine applications.

HL7 FHIR is a healthcare information exchange standard for EHR data.^54^ FHIR is being implemented in EHR systems worldwide, and provides the opportunity to deploy a more robust phenotypic representation in the context of recording the clinical encounter natively within the EHR. The GA4GH Clinical and Phenotypic Data Capture Work Stream has developed a FHIR Implementation Guide (IG) that focuses on elements of the Phenopacket schema that are most commonly used for rare-disease genomic diagnostics (see Web Resources). Although the GA4GH Phenopacket was not designed to explicitly mirror existing data schemas for cancer, it was designed to cover the majority of data elements in the mCODE FHIR IG^55^ and the International Cancer Genome Consortium ARGO Data dictionary^56^ and we envision that multiple different FHIR resources such as mCODE can be exported using the GA4GH Phenopacket schema, enabling software to be written that will take advantage of a unified data format. The Phenopacket FHIR IG is being further developed under the HL7 FHIR Vulcan program.^57^

The GA4GH has committed to coordinate its activities and future roadmaps with those of other standards development organizations (SDOs), including International Organization for Standardization (ISO) Technical Subcommittee for Genomics Informatics (ISO/TC215/SC1) and HL7 Clinical Genomics (CG). The ISO version of the Phenopacket schema documentation, titled “Genomics Informatics — Phenopackets: A Format for Phenotypic Data Exchange”, has been developed with input from both ISO and GA4GH stakeholders and aligned with the GA4GH Phenopacket schema. The final ISO balloting has been initiated, with publication planned for Spring 2022. This work will increase the availability of standardized phenotypic information and expand the collection of use cases to develop a standard relevant to genomics communities internationally.

### Applications of the Phenopacket schema

The Phenopacket schema was designed to support a number of use cases. Many of these use cases have been implemented and tested in the community, particularly in the field of rare disease diagnostics and biobanking, while others, such as EHR integration, are in the process of being implemented. These use cases and implementations are discussed below.

#### Rare-disease diagnostics

Over 10,000 rare diseases have been identified to date^58^, collectively affecting between 3.5% and 8% of the population,^59,60^ yet many patients experience a long diagnostic odyssey of 5-7 years.^44,58^ To date, each of the numerous tools for phenotype-driven genomic diagnostic support have used bespoke input formats for the VCF file representing variants called from exome or genome sequencing, phenotypic data (generally in the form of a list of HPO terms), and information about the pedigree. The Phenopacket provides a standard input format for these tools that will simplify computational analysis pipelines, especially if the steps in the pipeline include a comparison of the results of multiple tools. Exomiser,^29^ LIRICAL,^61^ Phen2Gene,^62^ and CADA^63^ can take Phenopackets as input files, and other analysis tools will soon accept phenotype data in Phenopacket format.

Multiple upstream data collection and management tools already support exporting patient profiles as phenopackets for downstream analysis and data sharing, including PhenoTips^64^ RD-Connect Genome-Phenome Analysis Platform (GPAP), Patient Archive in Australia and IRUD Exchange in Japan. PhenoTips can generate Phenopackets from patient or family records through a user interface or REST APIs, and includes de-identified demographic data, clinical phenotype, diagnoses, curated genetic findings, and pedigree data. Multiple projects, including the Children’s Mercy Research Institute’s Genomic Answers for Kids initiative, have adopted Phenopackets to help standardize data integration between PhenoTips and other systems. Projects such as EU funded Solve-RD and the European Joint Programme on Rare Diseases (EJP-RD) can generate Phenopackets for the data included in GPAP, which aims to facilitate diagnosis and novel gene discovery for clinical researchers.^65^ Phenopackets are used in Solve-RD to share phenotypic and other relevant clinical or genetic information (e.g., candidate or causative variants) between the consortium members, and are also deposited along the genomics data at the European Genome-phenome Archive (EGA) for long-term archival and controlled access. Besides being a successful instrument for data import/export between the project’s databases, Phenopackets have also proved to be useful for data analysis, such as clustering patients based on their phenotypic similarity. Solve-RD has so far generated phenopackets from 11,349 individuals; this number is expected to increase soon to over 19,000.^66^

#### European Joint Programme on Rare Diseases (EJP RD)

The EJP RD is developing a platform for federated discovery across rare diseases following the ‘data visiting’ strategy.^67,68^ The EJP RD is a GA4GH driver project, and several activities are making use of Phenopackets. On the topic of patient registries, the 24 involved European Reference Networks (ERNs) are working on applying the FAIR principles^41^ for the virtual data integration of different relevant resources in the platform. A FAIRification toolkit for the data stewards is under development to provide ready to use standards, where the Phenopacket schema is one of the planned clinical standards to be implemented. To improve the interoperability of Phenopackets within the FAIR platform, the project is providing *Semantic Phenopackets* based on Semantic Web standards and ontologies from the Open Biological and Biomedical Ontology (OBO) Foundry.^69,70^ The first release of this ontological modelling work is driven by the CAKUT (Congenital Anomalies of the Kidneys and Urinary Tract) rare disease registries use case^71^ Although at an early stage, the ability to seamlessly aggregate and integrate data from multiple registries or databases has huge potential benefits in terms of increasing the size of patient numbers for epidemiological analysis and facilitating the integration of distributed patient data into semantic knowledge graphs. To achieve full interoperability of ERNs rare disease patient registries with Phenopackets, the *Semantic Phenopackets* developed for the EJP RD virtual research platform will be further extended to include a full representation for all the elements of the GA4GH standard.

#### BioSamples and the European Genome-phenome Archive (EGA)

The BioSamples database at EMBL-EBI provides a central hub for sample metadata storage and linkage to other EMBL-EBI resources.^72^ BioSamples links to EGA^73^ and has done additional curation of human disease samples targeted to supporting Phenopackets. Every sample in BioSamples (over 19.5 million as of November 2021) exposes a Phenopacket record. Users can, for example, get a phenopacket for every single COVID-19 sample (see URLs).

#### Biobanks

In Japan, the Agency for Medical Research and Development (AMED) nation-wide biobank network has been developed to connect 12 major biobanks including Biobank Japan, Tohoku Medical Megabank project, and National Center Biobank Network. Together, the network stores over 855,181 biospecimens and 203,741 genomic data by 421,861 donors that are made available through a common query system exposing all phenotype data as phenopackets for downloading.

#### Journals

Published case reports contain data of immense value for characterizing and quantifying both new and existing conditions. However, these data are usually inaccessible for computational use, because they are represented in free-text or tabular formats designed for human readability. Even if the data are labeled using standardized ontology terms, there is no standardized format that can be used to disseminate the researchers’ findings to the wider community. We envision that authors of articles that include descriptions of patients (case reports) could submit representations of the patients as Phenopackets at the time of manuscript submission to journals. This model is analogous to the structural biology field, where biological three-dimensional structures are required to be deposited with the Worldwide PDB (wwPDB) organization before publication. The wwPDB remains responsible for curating, storing and making structure data accessible, which maximizes the impact and utility of any work, both to the scientific community and from a funding perspective. Like with the structural biology community, a broader solution to the problem of providing access to structured phenotypic data in journal articles would be a collaboration between journals and databases, such as the EGA and the BioSamples database, to provide accessions and help authors deposit their data in a FAIR way.

#### Patients and registries

From the patient perspective, improved phenotype data sharing offers individuals and families affected by a disease the opportunity to share readily observable data through patient-centered phenotyping approaches. The recent translation of the clinically focused HPO into layperson terms^52^ and ongoing Indigenous language translations of the HPO under the Lyfe Languages initiative will facilitate patient-driven phenotyping and subsequent matching and discovery of additional patients. The ability to use Phenopackets is also incorporated in the new architecture of the Western Australian (WA) Register of Developmental Anomalies, which includes two registers, the WA Birth Defects Register and the WA Cerebral Palsy Register, and is connected to multiple health data sets and is part of the data linkage ecosystem in the WA Health Department.

### Configuring Phenopackets

Because of the broad range of intended use cases and the large number of terminologies and ontologies in use by different communities, the Phenopacket schema is intentionally flexible with respect to which elements are required and which terminologies or ontologies must be used. Nonetheless, a given hospital, project, or research consortium may wish to apply different constraints. For instance, a Mendelian genetics consortium might stipulate the use of HPO terms to describe phenotypic abnormalities, and a cancer genomics consortium might require that each Phenopacket have a biosample for a tumor biopsy in which NCIT terms are used to describe the phenotypic features and other data.

The Phenopacket schema itself has minimal requirements and most of the top-level fields are optional. The requirements are intended to enforce that elements have sufficient data to be unambiguous without requiring information that may not always be available. For instance, if the *Quantity* element is used, it must have the *unit* and *value* fields, but the *referenceRange* field is optional. The requirements for each element are defined in the online documentation. We have implemented a Java library and command-line application that validates Phenopackets using a JSON schema to enforce these constraints. The library, phenopacket-validator (see Web Resources), enables users to specify additional project- or consortium-specific constraints and to enforce validity of ontology terms used (examples are provided in the GitHub repository).

## Outlook

Our goal is to provide a computable, well-structured representation of an individual’s medically relevant data to serve a wide community of patients and clinicians, scientists, and software developers. A schema analogous to the VCF standard for genetic variation has been lacking for clinical data such as phenotypic features, disease diagnoses, and treatments. We have therefore developed the Phenopacket schema within the broad community of GA4GH to fill this need. Just as VCF is not currently able to represent all types of genomic variation, the Phenopacket schema does not cover all potentially relevant types of clinical information. The Phenopacket schema was designed to be easily integrated with other schemas that use the protobuf or JSON frameworks, and we envision future work to integrate the Phenopacket schema with standards to represent other areas including population background (‘race’ or ‘ethnicity’), social determinants of health, and environmental exposures.

Software has become an essential resource for genomic medicine. A common experience of software developers is that a substantial amount of time is spent on reformatting data to correspond with the input format demanded by analysis software, or in the development of bespoke data schemes for new research projects. We hope that the Phenopacket schema will encourage the development of a collection of software for the analysis of genomic data in the context of clinical information that will accelerate innovation and discovery.

While the need for standardized computable and comparable clinical and genomic information is the basis for the development and anticipated implementation of Phenopackets, this does not replace a clear, concise, and nuanced narrative case report that describes the patient and the course of the disease over time. We strongly encourage the continued inclusion of case reports in addition to phenopackets in future publications.

Genomic data will become ever more important in translational research and clinical care in the coming years and decades. The Phenopacket schema represents a standard for capturing clinical data and integrating it with genomic data that will help to obtain the maximal utility of this data for understanding disease and developing precision medicine approaches to therapy.

## Data Availability

All data produced are available online at https://github.com/phenopackets/phenopacket-schema

## Web Resources

### Core Phenopacket resources

Phenopacket schema source code: https://github.com/phenopackets/phenopacket-schema

henopacket schema documentation: https://phenopacket-schema.readthedocs.io/

Phenopacket validator: https://github.com/phenopackets/phenopacket-validator

Phenopacket tools: https://github.com/phenopackets/phenopacket-tools

### Related standards

GA4GH Beacon project: https://beacon-project.io/

GA4GH Phenopacket FHIR implementation guide: https://github.com/phenopackets/core-ig

A4GH Pedigree standard: https://github.com/GA4GH-Pedigree-Standard/pedigree

GA4GH Variation Representation Specification (VRS): vrs.ga4gh.org

VRS Added Tools for Interoperable Loquacious Exchange (VRSATILE): vrsatile.readthedocs.io

Phenopacket RDF model: https://github.com/LUMC-BioSemantics/phenopackets-rdf-schema/wiki

Genomics Informatics — Phenopackets: A Format for Phenotypic Data Exchange (ISO): https://www.iso.org/standard/79991.html

### Databases using Phenopackets

The European Joint Programme on Rare Diseases (EJP RD): https://www.ejprarediseases.org/

BioSamples: https://www.ebi.ac.uk/biosamples (Search for NCBITaxon_2697049 to see BioSamples related to Severe acute respiratory syndrome coronavirus 2 or SAMN17024786 for a specific example).

AMED Biobank Network : http://biobank-network.jp/.

## Acknowledgements

The authors gratefully acknowledge insight and feedback from Marian H. Adly, Orion J. Buske, Pier Luigi Buttigieg, Nour Gazzaz, Janine Lewis, Manuel Posada de la Paz and Maria Taboada

## Contributions

PNR contributed clinical data model expertise, clinical subject matter expertise, data analysis, data curation, data integration, data quality assurance, funding acquisition, manuscript drafting, critical revision of the manuscript for important intellectual content, software engineering, statistical analysis. JSB contributed clinical data model expertise, clinical subject matter expertise, critical revision of the manuscript for important intellectual content. HH collaborated on data integration, data quality assurance, funding acquisition, manuscript drafting, critical revision of the manuscript for important intellectual content, software engineering. PB is acknowledged for biological subject matter expertise. GIS collaborated on biological subject matter expertise, clinical subject matter expertise, data curation, data integration, database / information systems admin, funding acquisition, governance, marketing and communications, project evaluation. SK contributed clinical data model expertise, data integration. CGC contributed clinical data model expertise, funding acquisition, governance, critical revision of the manuscript for important intellectual content, project management. KCL contributed funding acquisition, critical revision of the manuscript for important intellectual content. PMK contributed biological subject matter expertise, clinical data model expertise, clinical subject matter expertise. MB contributed biological subject matter expertise, clinical data model expertise, database / information systems admin, manuscript drafting, project evaluation, software engineering. AHW contributed clinical data model expertise, data integration, manuscript drafting, software engineering. CFB collaborated on critical revision of the manuscript for important intellectual content. CJM contributed software engineering. AM-J contributed clinical data model expertise, software engineering. MT is acknowledged for. MP is acknowledged for biological subject matter expertise, clinical data model expertise, clinical subject matter expertise, data analysis, data curation, data integration, data quality assurance, data visualization, database / information systems admin, funding acquisition, governance, manuscript drafting, critical revision of the manuscript for important intellectual content, marketing and communications, project evaluation, project management, regulatory oversight / admin, software engineering, statistical analysis. DS contributed data analysis, data quality assurance, funding acquisition, project evaluation. ST contributed clinical data model expertise, clinical subject matter expertise, data integration. NAV contributed data curation. BPC collaborated on clinical data model expertise, clinical subject matter expertise, critical revision of the manuscript for important intellectual content. MD collaborated on biological subject matter expertise, clinical data model expertise, data integration, data quality assurance, manuscript drafting, critical revision of the manuscript for important intellectual content. HC collaborated on clinical subject matter expertise, data analysis, data curation, data integration, critical revision of the manuscript for important intellectual content. GSB contributed clinical subject matter expertise, funding acquisition, manuscript drafting, marketing and communications. LB collaborated on data analysis, data integration, software engineering. MC contributed biological subject matter expertise, clinical data model expertise, data curation, data integration, data quality assurance, data visualization, database / information systems admin, critical revision of the manuscript for important intellectual content, software engineering. TG contributed data integration, software engineering. VAK collaborated on biological subject matter expertise, clinical data model expertise, clinical subject matter expertise. PIS collaborated on clinical data model expertise, clinical subject matter expertise, data curation, data quality assurance, critical revision of the manuscript for important intellectual content. AH contributed clinical data model expertise, clinical subject matter expertise. PNS contributed clinical data model expertise, manuscript drafting, critical revision of the manuscript for important intellectual content. AZ collaborated on clinical data model expertise, clinical subject matter expertise, critical revision of the manuscript for important intellectual content. NG is acknowledged for critical revision of the manuscript for important intellectual content. MAH contributed biological subject matter expertise, clinical data model expertise, data curation, funding acquisition, governance, manuscript drafting, critical revision of the manuscript for important intellectual content, marketing and communications, project management. NLH collaborated on funding acquisition, governance, critical revision of the manuscript for important intellectual content, project management. NP contributed database / information systems admin, critical revision of the manuscript for important intellectual content. NQ-R contributed clinical data model expertise, manuscript drafting, critical revision of the manuscript for important intellectual content. MCM-T contributed governance, project evaluation, project management. CW contributed data integration. AK contributed clinical data model expertise, data analysis, governance, critical revision of the manuscript for important intellectual content. DPH collaborated on clinical data model expertise. OE contributed biological subject matter expertise, clinical subject matter expertise. HL collaborated on biological subject matter expertise, clinical subject matter expertise, funding acquisition, governance. RT collaborated on biological subject matter expertise, clinical data model expertise, clinical subject matter expertise, data curation, funding acquisition. ARM collaborated on clinical data model expertise. JL is acknowledged for biological subject matter expertise, manuscript drafting. JOBJ contributed clinical data model expertise, data analysis, data integration, data quality assurance, manuscript drafting, marketing and communications, project management, software engineering. RS contributed manuscript drafting, critical revision of the manuscript for important intellectual content, software engineering. OJB is acknowledged for critical revision of the manuscript for important intellectual content, marketing and communications. EMS contributed clinical data model expertise, clinical subject matter expertise, data curation, data integration, data quality assurance, critical revision of the manuscript for important intellectual content. MA collaborated on biological subject matter expertise, critical revision of the manuscript for important intellectual content. JS contributed data curation, critical revision of the manuscript for important intellectual content. MHA is acknowledged for clinical data model expertise, clinical subject matter expertise, data analysis, data integration, data quality assurance, database / information systems admin, governance, manuscript drafting, critical revision of the manuscript for important intellectual content, statistical analysis. LDS contributed manuscript drafting. JAM contributed data visualization, funding acquisition, manuscript drafting, critical revision of the manuscript for important intellectual content. DD contributed software engineering. MAG contributed data integration, software engineering. HL contributed biological subject matter expertise, critical revision of the manuscript for important intellectual content. SO contributed clinical data model expertise, manuscript drafting, project evaluation. TJC contributed clinical data model expertise, data analysis, data integration. AS contributed clinical data model expertise, data integration, software engineering. JLW contributed clinical subject matter expertise. DP contributed software engineering. LM contributed software engineering. AP contributed software engineering. SB contributed manuscript drafting, software engineering. MR contributed software engineering. RK contributed data integration, software engineering. BJL contributed clinical subject matter expertise. RRF contributed biological subject matter expertise, clinical data model expertise, data integration, funding acquisition, governance.

